# The effects of border control and quarantine measures on global spread of COVID-19

**DOI:** 10.1101/2020.03.13.20035261

**Authors:** M. Pear Hossain, Alvin Junus, Xiaolin Zhu, Pengfei Jia, Tzai-Hung Wen, Dirk Pfeiffer, Hsiang-Yu Yuan

## Abstract

The rapid expansion of coronavirus (COVID-19) has been observed in many parts of the world. Many newly reported cases of this new coronavirus during early outbreak phases have been associated with travel history from an epidemic region (identified as imported cases). For those cases without travel history, the risk of wider spreads through community contact is even higher. However, most population models assume a homogeneous infected population without considering that the imported and secondary cases contracted by the imported cases can pose a different risk to community spread.

We have developed an “easy-to-use” mathematical framework extending from a meta-population model embedding city-to-city connections to stratify the dynamics of transmission waves caused by imported, secondary, and others from an outbreak source region when control measures are considered. Using the dynamics of the secondary cases, we are able to determine the probability of community spread.

Using the top 10 visiting cities from Wuhan in China as an example, we first demonstrated that the arrival time and the dynamics of the outbreaks at these cities can be successfully predicted under the reproductive number *R*_0_ = 2.92 and latent period *τ* = 5.2 days. Next, we showed that although control measures can gain extra 32.5 and 44.0 days in arrival time through a high intensive border control measure and a shorter time to quarantine under a low *R*_0_ (1.4), if the *R*_0_ is higher (2.92), only 10 extra days can be gained for each of the same measures. This suggests the importance of lowering the incidence at source regions together with infectious disease control measures in susceptible regions. The study allows us to assess the effects of border control and quarantine measures on the emergence and the global spread in a fully connected world using the dynamics of the secondary cases.

## Introduction

On 31 December 2019, the World Health Organization (WHO) was alerted to several cases of pneumonia infections in Wuhan City, Hubei Province of China [1]. The cause of the pneumonia was later identified as a novel coronavirus (COVID-19) genetically closely related to the Middle Eastern Respiratory Syndrome virus (MERS-CoV) and the Severe Acute Respiratory Syndrome virus (SARSCoV) [2]. The novel virus was able to establish human-to-human transmission [3] and caused high mortality and morbidity rates. Just after one month, more than a hundred confirmed cases have been reported all over the world outside China, not only in nearby countries like Japan, Korea, Singapore, but also in Europe and the Americas [4]. The WHO has since declared the outbreak an international emergency.

During the outbreak emergence, one of the most urgent public health tasks is to prevent the spread of the virus from an epidemic source region to other regions within a country or globally. Because a person who was infected can travel to another region and spread the virus, COVID-19 continued to pose a severe threat to other regions through transportation services. Many newly reported cases of this new coronavirus infection in other cities or countries before community spread have been associated with travel history from an epidemic source region or contact history to people from the region, referred as the imported cases and the secondary cases transmitted from the imported cases respectively. Once the secondary cases continue to transmit to more local cases, infection chain is established in the community, community spread begins subsequently [5].

The importance of prediction of infectious diseases based on transportation network information has already been highlighted in many previous studies [6, 7]. The probability of emergence and the arrival time of the emergence can be estimated using different approaches through model simulation with intensive computation or analytical expressions based on complex network structure with Poisson processes [8, 9, 10, 11]. However, due to intensive simulation or complex structure, these studies may either provide limited insights into public health or may be difficult to be implemented. A swift response in public health decisions is required for society’s infectious disease emergency preparedness. From infectious disease control perspective, the major questions here are whether we can i) assess the risk for COVID-19 to spread to other cities or countries to cause community spread; and ii) evaluate the effects of infectious disease control by cessation of population movement (e.g., lockdown, border control, or quarantine measures) on the outbreak spreading. Thus, an easy-to-use mathematical formula that can be embedded in a classical meta-population model to evaluate the risk of community spread and the effects of control measures is needed.

The increasing number of secondary cases indicate a significant risk of community spread. Using the Severe Acute Respiratory Syndrome (SARS) outbreak as an example, the progression of the SARS outbreak during a given time was generally following this order. First, people migrated from a city with the outbreak. Thus, infected cases arrived in other cities or countries as imported cases; second, the imported cases began to infect those who had very close contacts with them, resulting in secondary cases [12]; third, local transmission began after infections occurred for people without contact history with the imported cases or travel history to the outbreak source [13]. After these processes occurred repeatedly in many regions, SARS eventually spread rapidly throughout the world via air travel. Normally the imported cases can be soon isolated or quarantined due to the strict control policy against COVID-19 spread. However, contact tracing and isolation of cases are more difficult to be performed on secondary cases than imported, especially when *R*_0_ is high [14]. To prevent community transmission to happen, early prediction of the cumulative secondary cases generated by the imported cases is extremely critical.

Although disease compartment models with meta-population are widely used with migration or transportation data, the complexity of using these models sometimes infuriate infectious disease modelers when an urgent response is needed because most of the meta-population models do not offer a simple way to estimate the dynamics of imported and secondary cases [13, 10]. How to produce the dynamics of these cases in a simple way when the incidence is still increasing at the source remains to be answered. In addition, in order to evaluate the impact of infectious disease control measures on virus spread, there is a need to estimate the probability of emergence and the arrival time of the emergence through secondary infections under different control measures.

In this study, we have built a meta-population model based on a classical SIR model coupled with a mobility matrix and mathematical expressions to understand the outbreak spreading dynamics stratified by imported, secondary and other local cases at different cities. We have generate mathematical expressions that can be embedded into the SIR model to estimate the outbreak potential at neighboring cities. These formulas allowed us to calculate the dynamics of the first wave (imported cases) and the second wave of transmission (secondary cases produced by the imported cases). Using the cumulative number of the secondary cases, we thus predicted the probability of outbreak emergence and evaluated border closure and quarantine measures against this nation-wide outbreak spreading. Gain time before outbreak emergence was predicted under different control measures with different *R*_0_ settings.

## Materials & Methods

### Meta-population model

Assuming the newly emergence of COVID-19 causes an outbreak at location *i*, during the emergence, the changes of the numbers of infectious cases *I*_*j*_ at a different location *j* can be determined using a simple Susceptible, Infected, and Recovered (SIR) meta-population model with a mobility matrix (contact mixing at the population level):

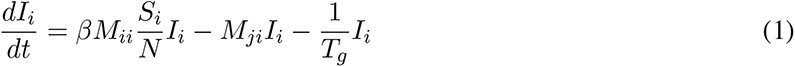

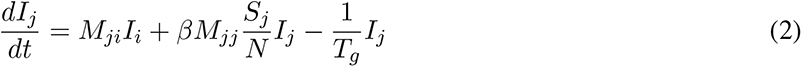

where *β* is the baseline transmission rate that can be estimated from 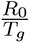, *R*_0_ is the basic reproductive number, *T*_*g*_ is the generation time, *M*_*ii*_ is the human mobility rate within the source location *i, M*_*ji*_ is the mobility rate from *i* to *j*, and *I*_*j*_ is the number of infected individuals at the location *j*. Our aim was to develop a meta-population model embedded with analytical expressions that can stratify the imported cases and the secondary cases produced by the imported cases along with other infected individuals with border control and quarantine measures. We modified this simple meta-population model by introducing the effect of border control and the quarantine. The mobility rate was multiplied by (1 *− c*), where *c* represents a border control measure. When *c* is higher up to 1, the mobility rate is reduced to zero. The infected cases were quarantined on average *T*_*qr*_ days after they are transmitted. After derivation (steps are described in later sections), the final model became:

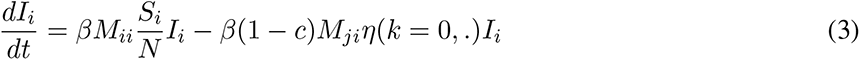

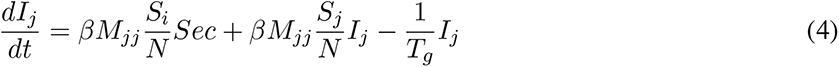

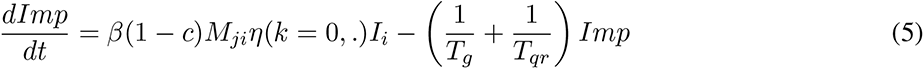

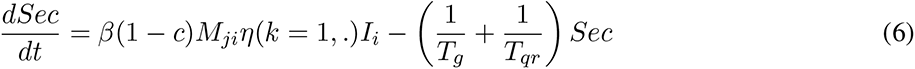

where *Imp* and *Sec* represent the imported and secondary cases produced by the imported cases (we will use secondary cases to denote this group in the remaining parts). We introduced an *η*(*k*,.) function to map the number of the infected at the source *i* to the imported and secondary cases at *j* given different border control and quarantine measures. The term *η*(*k* = 0,.) calculated the changes during first wave transmission (imported) and *η*(*k* = 1,.) calculated the changes during second wave transmission (secondary infected cases produced by the imported cases) under quarantine. The dot in *η*(*k* = 0,.) represents other epidemiological parameters.

During the early outbreak phase, because the susceptible population *S* was so close to the population size *N*, therefore, we assumed 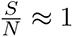. Because *M*_*ii*_ and *M*_*jj*_ are both near one (every day, more than 99.99% of individuals stay in the same location), we thus ignored the variables *M*_*ii*_ and *M*_*jj*_ to increase readability in the remaining sections.

### Calculating the arrival rate of imported cases

In order to calculate the imported cases, we assumed that infected cases could pass the border screening or move to another location only during their latent or incubation periods. We calculated the number of latent cases at time *s* by including a latent period *τ*. Therefore, the number of cases *I*_*i*_ in Eq(1) that were within the latent period at a specific time *s* were 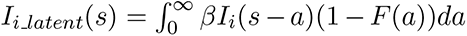, where 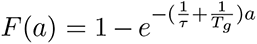 is the cumulative distribution function of latent cases that were transmitted *a* days ago but before recovery. The longer a latent period was, the more total imported cases were produced in a given duration. After replacing the number of infected cases by the latent cases, we obtained the rate of imported cases:

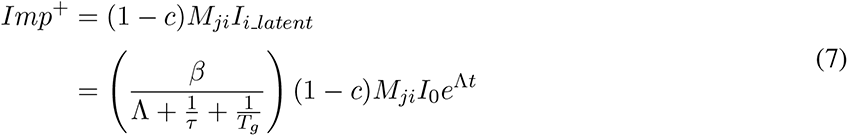

where Λ is the growth rate and can be calculated as 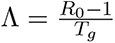 If all the infected cases can move to a different place as the latent period *τ* is quite long enough, the formula is reduced to (1 *− c*)*M*_*ji*_ *I*(*t*).

### Calculating numbers of imported and secondary cases

To calculate the number imported (*Imp*) and secondary cases (*Sec*), we had the following formulas now:

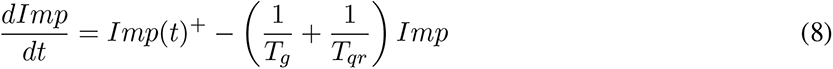

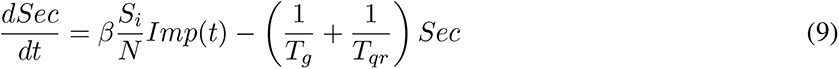

where *T*_*qr*_ is the time to quarantine. The number of cumulative imported cases can thus be derived during a certain period of time when the incidence is still exponentially increasing at the source location. Assuming 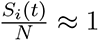, after solving the differential equation (detail derivation is available in the supplementary methods), the numbers of the imported cases and the secondary cases at time *t* were:

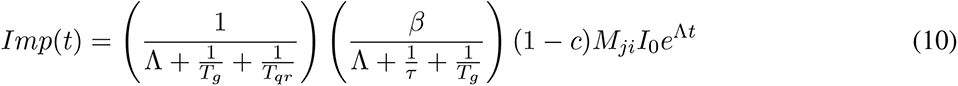

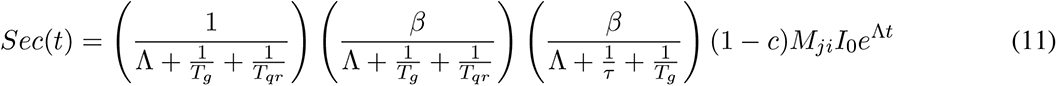

where *τ* is the latent period, Λ is the growth rate and 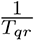 is the time to quarantine. Because of 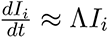, we have the infected number at *i* as a function of time *t*, that is *I*_*i*_ (*t*) = *I*_0_ *e*^Λ*t*^. If we set 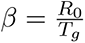, we obtained the infectious disease spreading control function given *k* transmission waves before the community transmission:

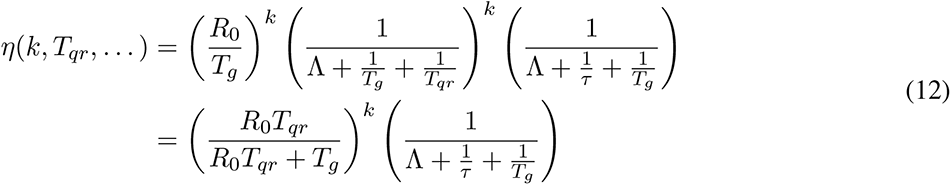

The formula was simplified after we replaced 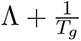 by 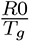 Under this notation, secondary infection cases can simply be calculated as *η*(*k* = 2,*⃛*) multiplied by the (1 *− c*)*M*_*ji*_ *I*_0_ *e*^Λ*t*^ :

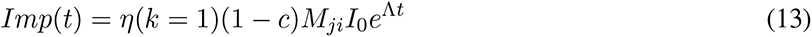

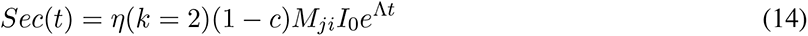

Consequently, the cumulative number of imported and secondary cases at time *t* under latent period were:

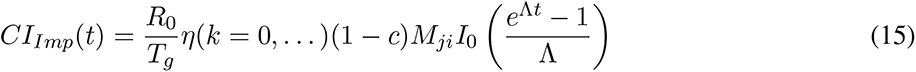

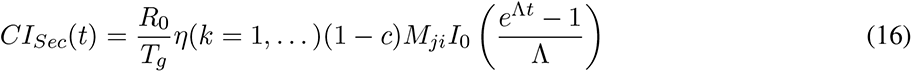

Because before the outbreak occurred (or before community spread) at a location *j*, infections only happened when transmission events occurred between the imported cases and the susceptible individuals at *j* (we call it the second wave of transmission), it is essential to estimate the cumulative number of the imported cases (*CI*_*Imp*_ ; the first wave of transmission) and the cumulative secondary cases transmitted from those imported cases (*CI*_*Sec*_ ; the second wave of transmission). We were able to deduce tertiary cases but the number will be relatively small comparing to the imported and the secondary.

Thus, we have a simple formula to predict certain important infection numbers before an outbreak emerges in a specific location using three epidemiological parameters reproductive number, generation time, incubation time and time to disease detection after onset along with a contact matrix. This framework provides a more generalized expression to estimate the cumulative imported cases (first wave of transmission) and the cumulative secondary infected cases generated by the imported cases (second wave of transmission) at connected cities or locations during a certain period of time when the incidence is exponentially increasing at the source.

### Constructing contact matrix

Airline passenger data were collected from the International Air Transport Association (IATA) database. We collected the actual passenger data for top 10 visiting cities leaving from Wuhan Tianhe International Airport before the lockdown of Wuhan city from 30 December to 20 January, 2020. Note that we did not have data for railroad and other forms of transport, and thus made an assumption that the total number of travelers is 4 times higher than that of air transport except for certain cities on Hainan island that have no road connections to Wuhan. We made this assumption because that the number of train passengers is few times higher than that of airline in China [15] and the results from a population migration database suggested a similar ratio [16]. The number of daily passengers between different cities were used to generate the mobility rate *M*_*ji*_ between locations *i* and *j*. For example, we divided a daily passenger number by total population size in Wuhan to represent the contact rate between Wuhan city and any other connected city *j*. We denote *i* = 0 as the index for the source city. Therefore, *M*_00_ = 1 is the base level of contact rate within the source city. *j* is a number to represent the index of the top visiting cities from first to last following the order in the list. Because the estimated number of people leaving from Wuhan to the first visiting city (Beijing) is 10793.5 per day, given that the population size of Wuhan is about 11 million, the daily percentage of people leaving to the 1st visiting city can be approximated to 0.001. Thus, we have *M*_10_ = 0.001. We used the same approach to determine *M*_*j*0_ for *j* = 2, 3, …, 10.

### Determining outbreak potential

Next we considered the outbreak potential, defined as the probability of outbreak emergence given the number of cumulative cases. At the initial stage, if there were *n* infected individuals, the chance of the viruses to cause an outbreak is 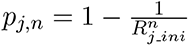 We determined a critical threshold number *v* to be 8 and set *p*_*j,v*_ = 50% after we compared the trends of the top 10 visiting cities. 50% of the cities demonstrated rapid growth of the numbers of infected cases once the numbers reached or near the threshold. We thus obtained the effective reproductive number *R*_*j ini*_ = 1.0905 to represent the transmissibility during the second wave of transmission at location *j*. Note that this number represents the epidemic growth under control measures before community spread. *R*_*j ini*_ indicates the average number of transmissions that are generated from the secondary cases (*Sec*) before the community spread. Because the nation-wide alert has already been received at different cities after 31 December, 2019, and many infectious disease control measures have been implemented, *R*_*j ini*_ was expected to be lower than *R*_0_. Given the *R*_*j ini*_ number, we used the cumulative number of secondary cases *CI*_*Sec*_ at the location *j* to calculate the probability of outbreak emergence as 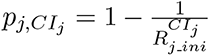 (to simplify the notation, the *CI*_*j*_ is used to represent *CI*_*Sec*_). We defined the critical arrival time such that the probability of outbreak emergence *p*_*j,CI*_ *j* was larger than 50%.

## Results

### Model of outbreak spreading

The cities that most Wuhan citizens moved to in the first month after the outbreak of the COVID-19 was reported in Wuhan posed a higher potential of outbreak emergence. We collected airline passenger data during 22 days after outbreak emerged between 30 December to 20 January, before the lockdown of Wuhan was implemented on 23 January, 2020. The daily airline passenger number from Wuhan to the top 10 visiting cities were extracted with the 1779.9 persons on average (Figure 1A). We estimated the total passengers leaving out from Wuhan using a human migration map airline passengers. We obtained the estimated numbers of total passengers given the proportion of airline passenger was about 23% of all transportation in early January [16]. An increasing number of the confirmed cases was observed among the top visiting cities, including Beijing, Shanghai, and Guangzhou (Figure 1B).

**Figure 1.**
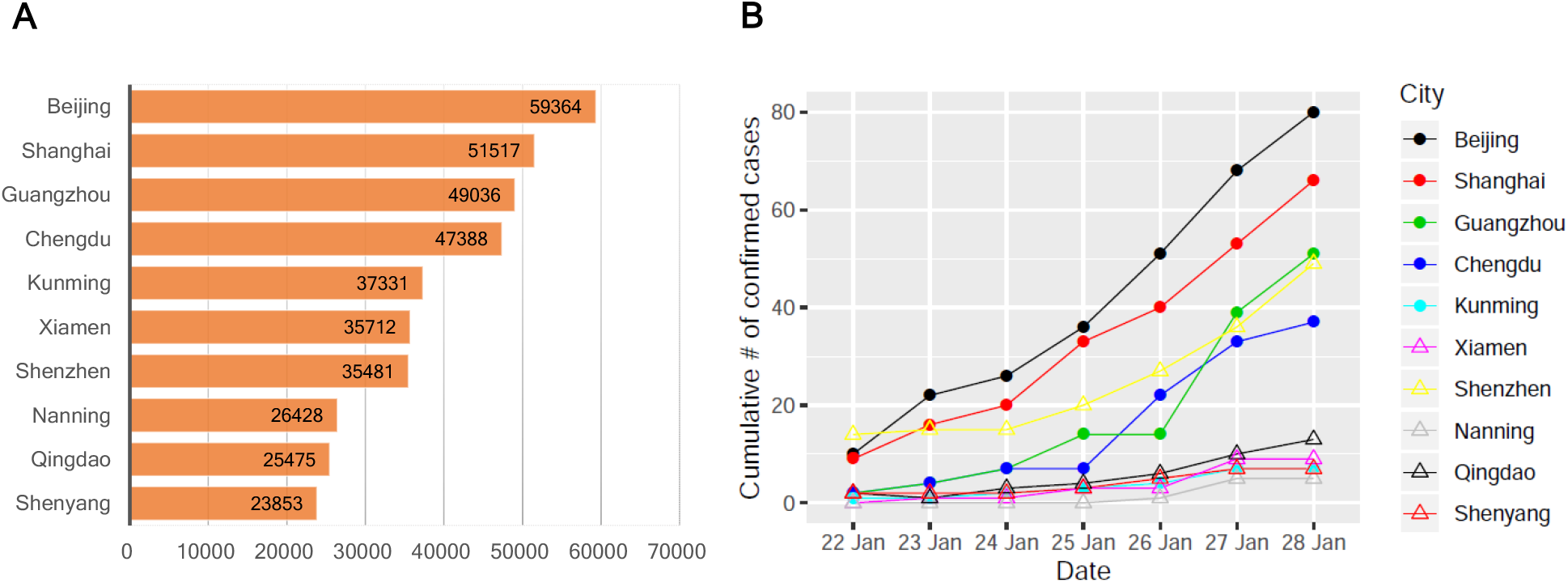
Statistics of airline passenger from Wuhan Tianhe International Airport and COVID-19 confirmed cases. (A) Number of airline passengers from Wuhan to top ten visiting cities between 30 December, 2019 to 20 January, 2020. (B) Number of confirmed cases at top ten visiting cities between 22 January to 28 January, 2020.

The community spread began as the number of the imported cases and the associated secondary cases generated by the imported cases accumulated to a certain number (Figure 2A). Thus, the increasing number of the imported cases can be correlated to the outbreak spreads through the number of departure passenger data. We developed a mathematical framework deriving from a standard meta-population model coupling with a migration matrix and incubation time with different control measures (Figure 2B) to calculate the number of the total imported cases and the cumulative secondary cases.

**Figure 2.**
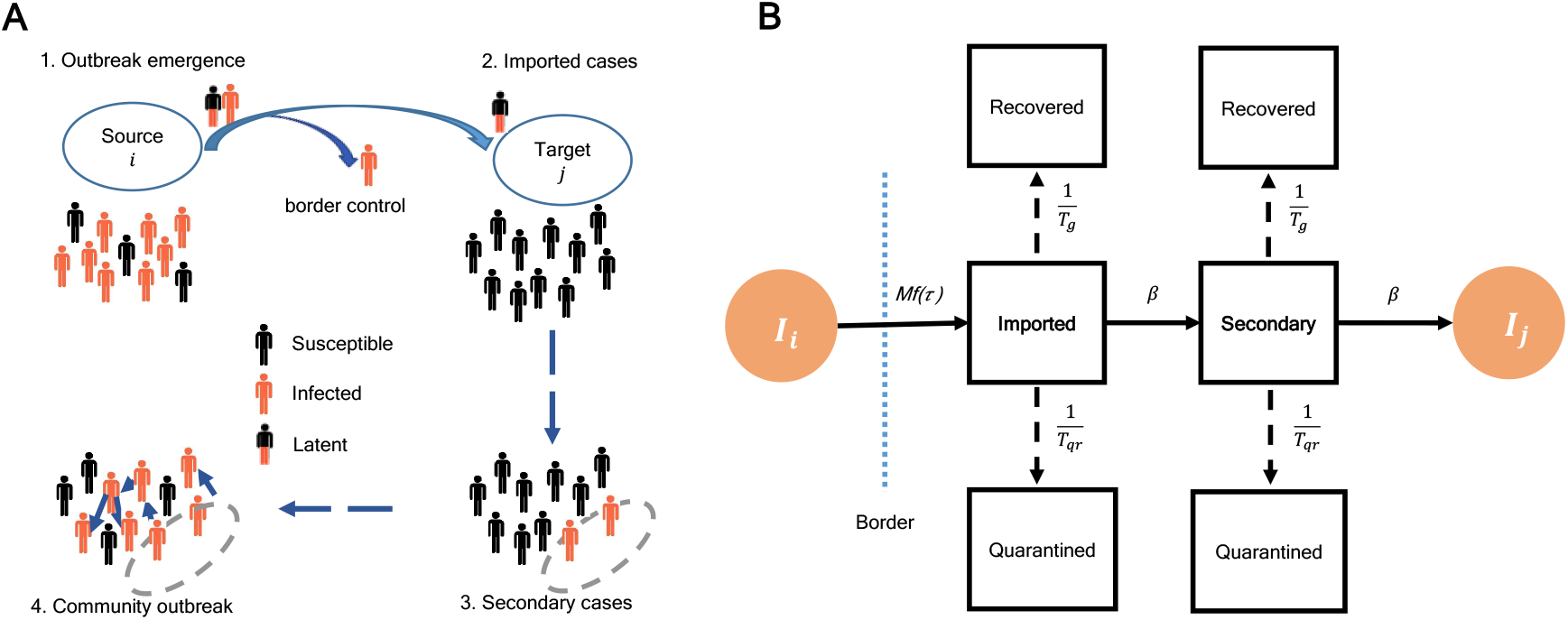
COVID-19 outbreak spread from source to community transmission. (A) Outbreak progression from source to community spread. The imported cases arrive after passengers passed the border control. The secondary cases produced by the imported cases eventually cause the community outbreak. The community outbreak starts after *t*. (B) Mathematical model framework for COVID-19 estimates of secondary cases. *I*_*i*_ and *I*_*j*_ represent number of infected cases in a source location *i* and a target location *j* respectively. *M* is the mobility rate and *f* (*τ*) is a function of incubation time that represent the percentage of infected cases that can pass the border (dashed line). *T*_*g*_ and *T*_*qr*_ are generation time and time to quarantine respectively. *β* estimates the ratio 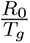, where *R*_0_ basic reproductive rate.

### Outbreak spreads in Mainland China

Outbreak spreads in Mainland China were reconstructed and compared with different transmissibilities and latent periods. We first calculated the cumulative number of secondary infected cases produced by imported cases among top 10 visiting cities from Wuhan under a scenario corresponding to an *R*_0_ of 2.92 [18], a generation time of 8.4 days [19], and an incubation time (for defining latent period) of 5.2 days for the outbreak in Wuhan city. The migration matrix *M* was constructed using the average of the airline passengers data during 22 days in January among these cities (*materials and methods*). The cumulative number of secondary infected individuals generated by the imported cases moving from Wuhan was calculated (Figure 3A). A threshold number of cases *v* = 8 was used to indicate a higher than 50% probability of community spread will occur if the cumulative number of secondary cases is over that threshold.

**Figure 3.**
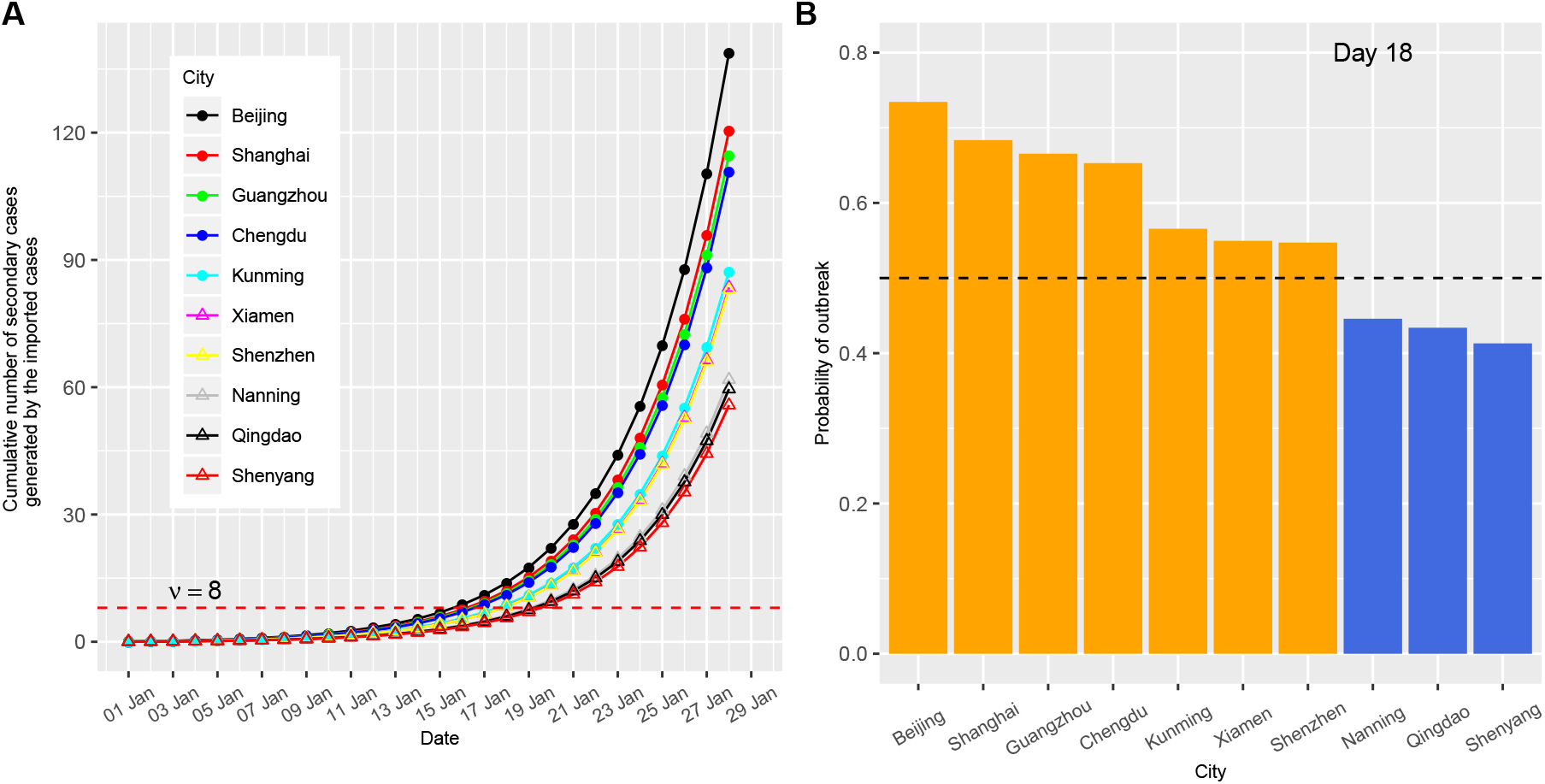
Outbreak potential estimated from the secondary cases contacted by imported cases. A higher *R*_0_ = 2.92 scenario with incubation time *τ* = 5.2 days and time to quarantine *T*_*qr*_ = 2 days were used. (A) Number of cumulative secondary cases generated by imported cases. The secondary infections are listed among the top 10 visiting cities from Wuhan. *v* = 8 is the critical threshold number; (B) Probability of outbreak emergence in different cities at mean arrival time (18 days).

The arrival times of outbreak emergence (defined as the time to reach above this threshold) for the top 10 cities were between 16 – 20 days (Figure 3A), with the average arrival time 18 days (corresponding to 18 January). On day 28, the secondary cases rise to 139 persons for the top city Beijing, which makes the total number of secondary cases among all the top ten visiting cities to 915.7 persons. The outbreak potentials of the cities were assessed on 18 January. 7 out of 10 cities have a probability of outbreak emergence larger than 50% (Figure 3B and *supplementary* Figure S8). Among those 7 cities, 5 of them had a high number of actual confirmed cases more than 36 on 28 January (10 days later), only 3 of them maintained low case numbers below 10. Taking into account that the actual lag of reporting time was about 10 days, the probability of outbreak emergence on 18 January indicates the level of the outbreak potential well (Figure 1B).

The predicted reporting delay was very close to the actual reporting lag. The average actual lag was calculated to be 10.30 days after counting the difference between the average date of onset peak (among 8 days with highest number of cases) and the average date of diagnosis peak (among 8 days with highest number of cases) shown in the recent report of 72314 cases from the Chinese Center for Disease Control and Prevention [20]. Note that we did not consider the long tail because no full longitudinal data were available yet. Parameter estimation of lag time using maximum likelihood approach to fit 10 cities together was 10.52 days (10.27-10.78; 95% confidence interval) for incubation time 5.2 days (*supplementary* Figure S5). Note that when we fitted each city individually, the reporting delay ranges from 8.0 to 18.3 days (*supplementary* Figure S6), with an average of 12.4 days. When only the top 5 cities with highest confirmed cases are considered, the reporting delay ranges from 8.0 to 11.2 days with an averages at 9.2 days. We did not rule out the possibility that the long delay time, such as 18.3 days, was due to actual delay of the outbreak emergence. Overall, the predicted cumulative number of both imported and secondary cases after adjusted by the reporting lag time of top ten visiting cities demonstrated a similar increasing trend in cumulative numbers of confirmed cases during each early emergence period (Figure 4 and *supplementary* Figure S7).

**Figure 4.**
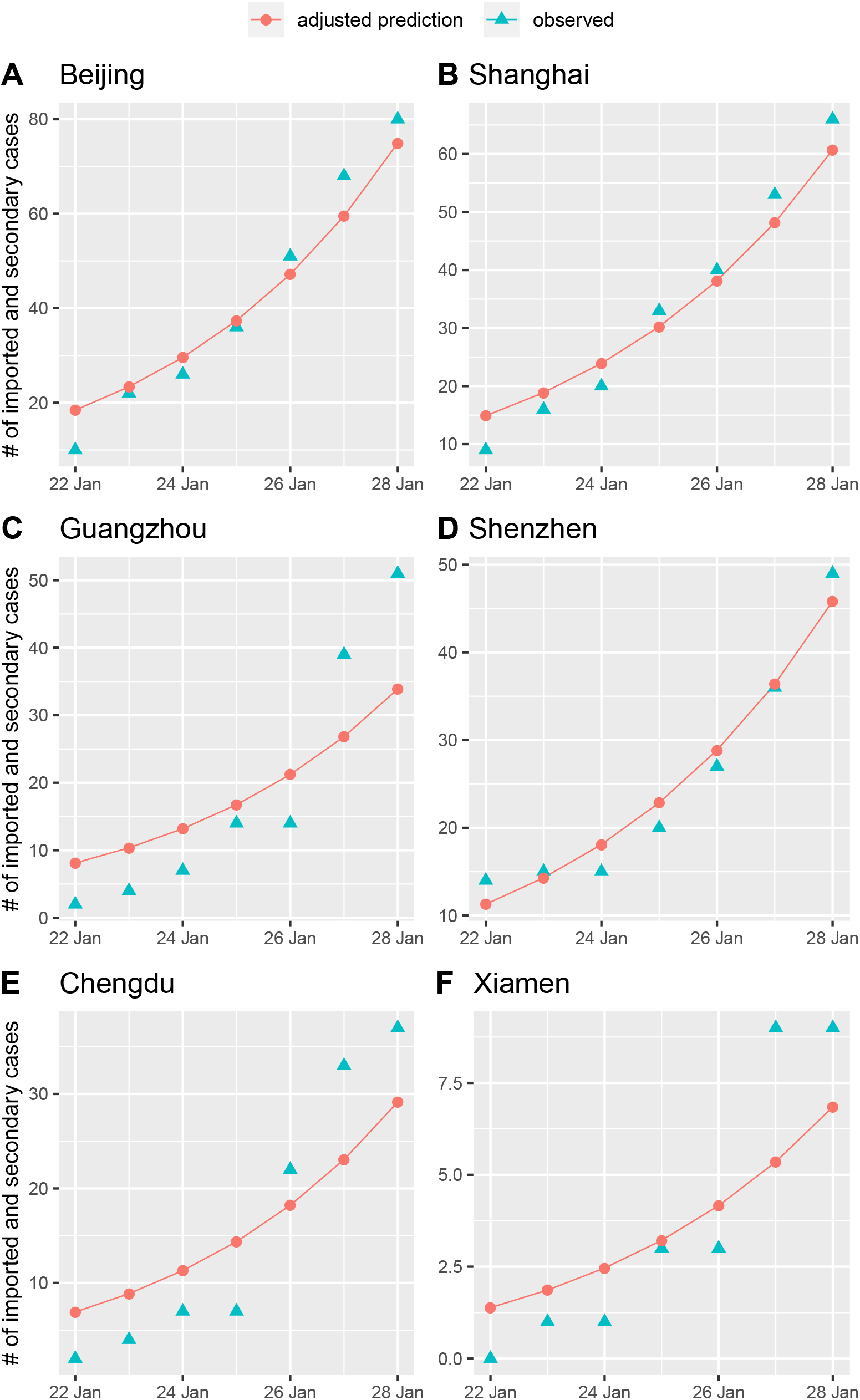
Observed number of confirmed cases and predicted number of imported and secondary cases. The predicted number of cases were adjusted by the reporting delay after using maximum likelihood estimation. The six cities that have the actual earliest arrival times are listed (four other remaining cities are given in the supplementary file).

In contrast, a longer latent period shortened the outbreak arrival time by allowing more secondary cases. Considering the same *R*_0_ setting but with incubation time 14 days, on day 28 the total number of secondary cases raised to 1179.8 persons for all the top ten visiting cities in which Beijing contributes 180 persons in the total secondary cases. Therefore, the longer latent period allowed 22% more secondary cases. The arrival time of outbreak emergence were 14-18 days (Figure 5A), which were 2 days earlier than using 5.2 days latent period. On day 18, all top ten cities except Shenyang have outbreak probability more than 50% (Figure 5B). The results confirmed that if latent period is long, more ill people can move to other cities.

**Figure 5.**
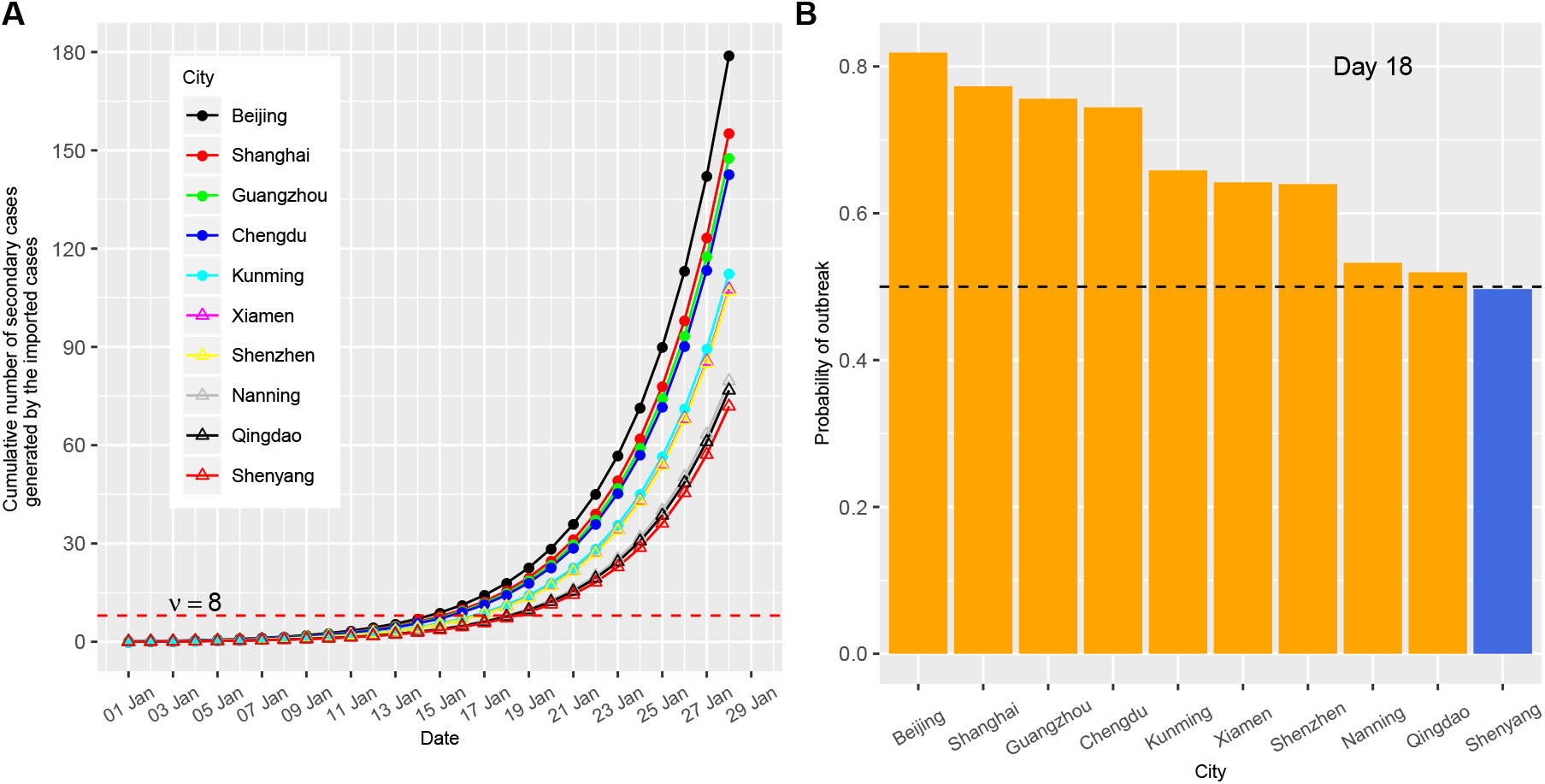
Outbreak potential estimated from the secondary cases contacted by imported cases. A higher *R*_0_ = 2.92 scenario with incubation time *τ* = 14 days and time to quarantine *T*_*qr*_ = 2 days were used. (A) Number of cumulative secondary cases generated by imported cases. The secondary infections are listed among the top 10 visiting cities from Wuhan. *v* = 8 is the critical threshold number; (B) Probability of outbreak emergence in different cities at critical time (18 days).

For each of the top ten cities, the arrival times of outbreak emergence for both 5.2 and 14 days latent periods were determined. The actual order of the arrival time for the top 4 cities, including Beijing, Shanghai, Guangzhou and Chengdu were successfully predicted (Table 1). A longer latent period produced a shorter arrival time.

**Table 1:** Actual and predicted arrival time of outbreak emergence at top ten connected cities in China. *R*_0_ = 2.92 with incubation time *τ* = 5.2 days and *τ* = 14 days were used. *CI*_*Sec*_ is the cumulative number of secondary infected cases generated by the imported cases. The actual arrival time of outbreak is defined as the the date when the number of cumulative cases is larger than the threshold number 8 and the number of newly reported cases is larger than 5. 9.2 days reporting lag between the date of onset and the date of diagnosis was estimated using the top five cities with most number of confirmed cases.

| City | Actual arrival time (day) | $\tau = 5.2$ | | | $\tau = 14$ | | |
| --- | --- | --- | --- | --- | --- | --- | --- |
| | | Predicted arrival time (day) | $CI_{Sec}$ at critical time (day 18) | Probability of outbreak at critical time (day 18) | Predicted arrival time (day) | $CI_{Sec}$ at critical time (day 18) | Probability of outbreak at critical time (day 18) |
| Beijing | <23 | 16 | 13.9 | 0.734 | 15 | 17.9 | 0.819 |
| Shanghai | <23 | 17 | 12.1 | 0.684 | 16 | 15.6 | 0.773 |
| Guangzhou | 25 | 17 | 11.5 | 0.666 | 16 | 14.8 | 0.756 |
| Chengdu | 26 | 17 | 11.1 | 0.653 | 16 | 14.3 | 0.744 |
| Kunming | 29 | 18 | 8.7 | 0.566 | 17 | 11.3 | 0.658 |
| Xiamen | 27 | 18 | 8.4 | 0.550 | 17 | 10.8 | 0.642 |
| Shenzhen | 25 | 18 | 8.3 | 0.547 | 17 | 10.7 | 0.640 |
| Nanning | >30 | 20 | 6.2 | 0.446 | 19 | 8.0 | 0.533 |
| Qingdao | >30 | 20 | 6.0 | 0.434 | 19 | 7.7 | 0.520 |
| Shenyang | >30 | 20 | 5.6 | 0.413 | 19 | 7.2 | 0.497 |

### Outbreak spreads with low transmissibility

Since *R*_0_ of COVID-19 was not fully known yet, we predicted the outbreak spreads using different *R*_0_ settings. In most cases, the effective reproductive number *R* may decrease after many control measures are conducted [13]. Therefore, in addition to *R*_0_ = 2.92, we made the same calculation under a low transmission setting with *R*_0_ = 1.4, which was the lower bound of World Health Organization’s estimate [21] and a medium transmission setting with *R*_0_ = 1.68, which was near the lower bound of *R*_0_ estimated by the MRC Centre for Global Infectious Disease Analysis at Imperial College London [22], to evaluate the arrival times of outbreak emergence among the top 10 visiting cities. Under these scenarios, following the suggestion from a recent study [23], we considered the initial infected number as 1000 on 31 December.

When *R*_0_ = 1.4, the cumulative secondary infected number was slowly linearly increasing and the top visiting city had 6.8 cases on 28 January, which was below the critical threshold line *v* = 8 (*supplementary* Figure S2). For each of the top ten cities, the arrival time at which the cumulative number of secondary cases larger than the critical threshold was determined (*supplementary* Table S1). Beijing, Shanghai, Guangzhou had the shortest required time periods. However, the arrival times of outbreak emergence in the above cities were between 31-34 days, corresponding to the end of January and the early of February, which were about more than 10 days later comparing to the actual reported data (*supplementary* Table S1). The mean arrival time of the top 10 visiting cities was 39 days, which was 21 days later than *R*_0_ = 2.92. With *R*_0_ = 1.68, the mean arrival time was 26.3 days. Given the reporting delay was about 10 days, we found that *R*_0_ = 2.92 with latent period 5.2 days gave a better prediction compared to other low *R*_0_ or long latent period settings.

### Impact assessment of border control and quarantine

We first evaluated the effects of border control measures by changing the control rate *c* to reduce the transportation between two cities under three different *R*_0_ settings. Border control or other special events, like lunar new year, can affect the traveling rates. Ideally, complete cessation of population movement between cities or isolation of every susceptible case from the source city can reduce transmission events most however it happens rarely. Then the extra arrival gain times of outbreak emergence comparing to a baseline setting using Beijing city were calculated.

Under a low *R*_0_ (1.4), the border control measure that reduced 90% of the passenger numbers gained extra 32.5 days of outbreak arrival time (Figure 6A). Under a medium *R*_0_ (1.68), we found that the effect of border control was weaker but still created 20.0 extra days under the same control level. However, under the high *R*_0_ (2.92), the effect on reducing the chance of outbreak emergence was very low, with only 10 extra days obtained.

**Figure 6.**
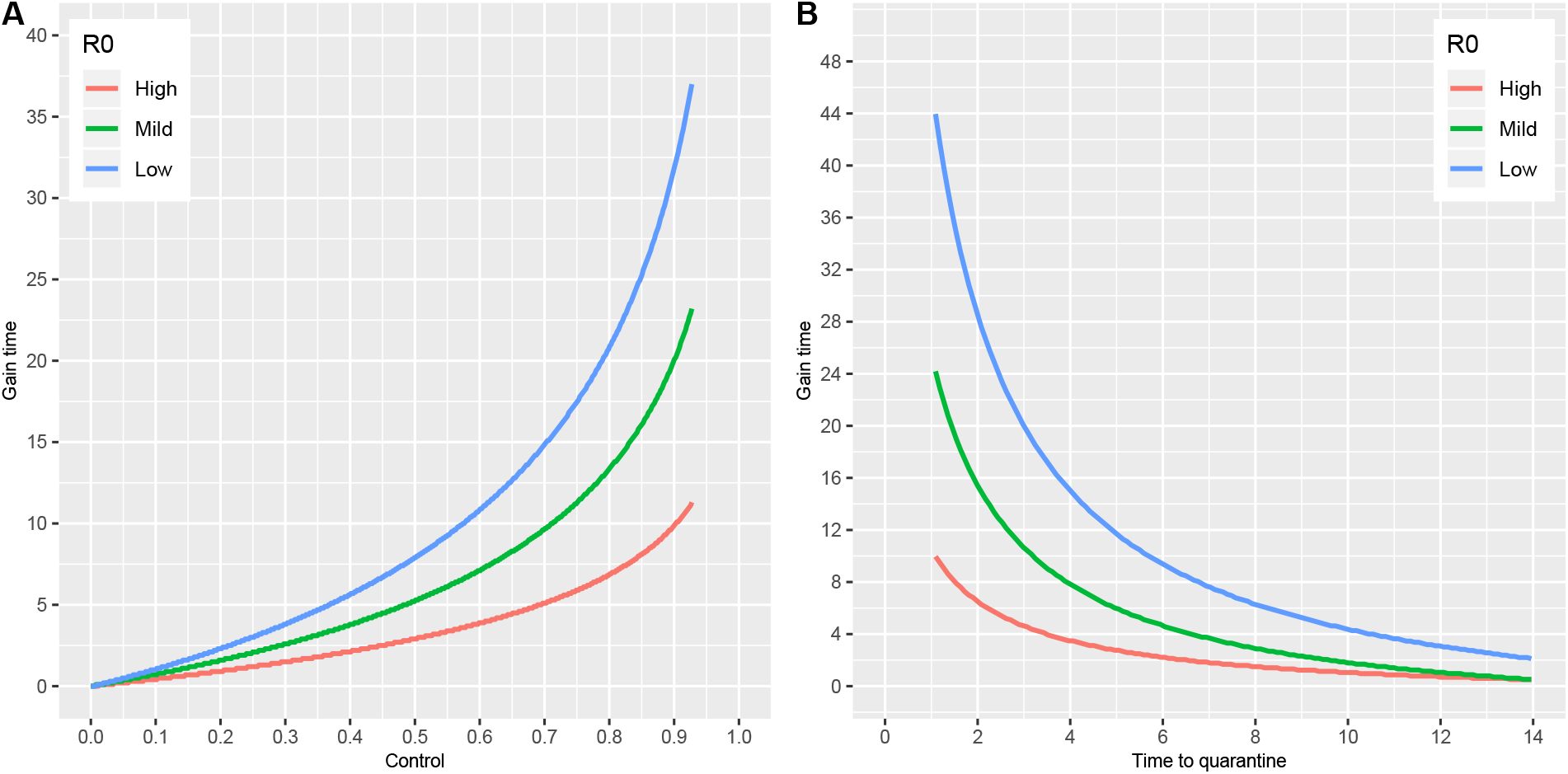
Assessment of border control and quarantine effects on outbreak arrivals. (A) Gain time of outbreak emergence by the rates of successful border control. The effects of border control on gain time under a low *R*_0_ (1.4), mild *R*_0_ (1.68), high *R*_0_ (2.92) were plotted in blue, green, and red colors. (B) Gain time of outbreak emergence by time to quarantined. The effects of border control on gain time under a low *R*_0_ (1.4), mild *R*_0_ (1.68), high *R*_0_ (2.92) were plotted using the same color codes as A. The passenger data of the top visiting city Beijing was used to generate the baseline arrival time. To get the gain time, the arrival time using different infectious disease control measures was calculated and was subtracted by the baseline arrival time.

Next, we calculated gain time by time to quarantine under different *R*_0_ settings. Under the low *R*_0_ (1.4), if the individual was quarantined immediately (happened in one day after the person became infectious), the gain time became as large as 44.0 days (Figure 6B). Under the medium *R*_0_ (1.68), we found that the quarantine effect was approximately half of the low *R*_0_ scenario (24.1 days) using the same time to quarantine. However, under the high *R*_0_ (2.92), the effect of quarantine was much lower, with only 10.0 days was gained.

## Discussion

The current epidemic marks the third time in 20 years that a member of the family of coronaviruses (CoVs) has caused an epidemic employing its zoonotic potential, for example, from bats [24]. The COVID-19 virus is able to establish between human-to-human transmission [3] and is currently spreading from Wuhan to many nearby cities and countries. It is hypothesized that the rate of transmission between different cities or countries is related to the number of people moving from different locations to Wuhan. About 2 weeks later after 31 December, using the number of cases detected outside China, it has been inferred that more than a thousand individuals (with an estimated mean 1723) had had an onset of symptoms by 12 January, 2020 [25].

The study demonstrated that after reporting delay was estimated, the dynamics of the outbreaks at connected cities can be successfully reconstructed using both the imported and the secondary cases. The result implies that all the connected (direct or indirect) countries are having a great risk of outbreak. The question is about when the outbreak will arrive [26] and how to delay the arrival time to have a better preparation. The challenge, however, is that we lack a simple and accurate tool for assessing outbreak emergence risk and subsequently the required levels of border control and quarantine measures to prevent additional outbreaks.

Until recently, although some studies have been done to predict the spreading of this new disease using air and other forms of transport information [6, 7, 27, 28, 29], none of studies were designed to estimate the dynamics of the imported and secondary cases. The benefit of stratifying the imported and secondary cases in a disease transmission model is to provide a risk assessment of community spread. Because most of the imported cases can be detected more easily under 14 days quarantine from the passengers coming from the epidemic source region, thus the risk of outbreak is not primarily linked to the number of the imported cases. However, secondary cases, without travel history to the epidemic source region, are more difficult to be identified or quarantined before disease onset and thus are more easily to become undetected cases in a community.

We evaluated the effectiveness of different infectious disease control on the secondary cases in order to estimate the arrival time of future community spread. Recent studies have shown the importance of modeling in infectious disease control [30]. A recent study has used a modeling approach to forecast the dynamics of outbreak spreading [27]. We developed an “easy-to-use” mathematical formula that are able to have an analytical solution of the first wave transmission (imported cases) and the second wave transmission (secondary cases generated by the imported cases). With these numbers, we are able to evaluate the impact of border control and quarantine measures. Surprisingly, under the higher *R*_0_ setting (2.92), the effect on obtaining 10 extra days requires an enhanced border control measure to reduce more than 90% of the passengers or a very efficient quarantine measure. The results suggest that if the epidemic growth at the source location is high, even a near full-scale border control without proper quarantine measure, will have only limited effects. The transmission waves can be treated as a branching process. However, instead of using the offspring variability to estimate the probability of extinction, we adopted a classical way to derive probability of extinction that was based on *R*_0_ or effective *R*.

We have learnt from the previous SARS outbreak that it is crucial to implement rapid infection control measures to limit the impact of epidemics, both in terms of preventing more casualties and shortening the epidemic period. Delaying the institution of control measures by 1 week would have nearly tripled the epidemic size and would have increased the expected epidemic duration by 4 weeks [31]. Previous study showed that control measures at international cross-borders and screening at borders are influential in mitigating the spread of infectious diseases [32] [33]. Cross-boarder screening system to prevent infectious disease outbreak is important but cannot successfully prevent ill persons during their latent period.

Full-scale border control measures to prevent the spread of the virus have been discussed in many countries in the world. At the same time, a lockdown of Wuhan city (border control from leaving out) has already been imposed. Here the model we constructed can be used to estimate the dynamics of imported and secondary cases using transportation data with different control measurements. The framework can be extended to multiple infected sources to multiple target cities without increasing the complexity of the computation dramatically. Hence, the model proposed in the current study could provide a risk assessment of COVID-19 global spreading in a highly connected world.

## Data Availability

.

## Acknowledgments

We thank Prof. Steven Riley from MRC Centre for Global Infectious Disease Analysis at Imperial College London to give valuable comments and thank Miss Xiaoyue Tan for her assistance in making maps used in the manuscript. We thank Prof. Mengsu (Michael) Yang, Mesfin Tsegaye from City University of Hong Kong and all the anonymous readers who have provided invaluable comments. The authors also acknowledge the support from the grants funded by City University of Hong Kong [#7200573 and #9610416] and the ministry of Science and Technology in Taiwan [MOST 108-2638-H-002-002-MY2].

## Author Contributions

HY and XZ designed the study. PJ participated in the data collection. HY, MPH, XZ, PJ and AJ analysed and interpreted the data. MPH, AJ and HY wrote the paper. Everyone reviewed, revised and edited the manuscript.

